# Cross-Disorder Machine Learning Uncovers Schizophrenia Risk Variants Predictive of Alzheimer’s Disease

**DOI:** 10.1101/2025.04.24.25326362

**Authors:** Taeho Jo, Carol S. Lim

## Abstract

Alzheimer’s disease (AD) and Schizophrenia (SCZ) exhibit overlapping clinical features and biological mechanisms, but the extent of their shared genetic etiology and the potential for cross-disorder risk prediction are not fully elucidated, with previous genetic studies yielding mixed results. This study investigated whether integrating statistically-selected SCZ-associated single nucleotide polymorphisms (SNPs), identified from large-scale GWAS, could enhance machine learning (ML)-based prediction of AD risk beyond models using only AD-associated SNPs. Utilizing summary statistics from large-scale GWAS for AD (N=1,126,563) and SCZ (N=320,404), alongside reference genotypes from the Alzheimer’s Disease Sequencing Project (ADSP, N=7,416 Non-Hispanic Whites), we employed an objective iterative ML framework evaluating distinct classification algorithms and adding prioritized SCZ SNPs to a baseline AD-SNP model. Integrating the top 50 SCZ SNPs significantly improved AD prediction accuracy in the best-performing ML model, increasing the area under the receiver operating characteristic curve (AUC) from 0.6032 (baseline) to 0.6400 (DeLong’s test p=0.0005). Contributing SCZ SNPs implicated shared biological pathways relevant to AD, including immune regulation/neuroinflammation, tau protein biology, and synaptic vesicle trafficking, while also revealing novel predictive variants warranting further investigation. These findings demonstrate that incorporating specific SCZ genetic risk variants can modestly but significantly enhance AD risk prediction, supporting meaningful genetic overlap and providing novel genetic targets for cross-disorder research, highlighting the potential of this approach for dissecting the complex genetic architecture of brain disorders.

## Introduction

Schizophrenia and Alzheimer’s disease (AD) are distinct neuropsychiatric disorders, both highly heritable (up to 80%)^1^ and characterized by cognitive impairment that differs in onset and progression. Schizophrenia typically manifests in adolescence or early adulthood, initially presenting as cognitive dysfunction long before psychosis emerges—historically described as “dementia praecox.”^2,3^ In contrast, AD is a progressive neurodegenerative disorder predominantly affecting older adults. Psychosis occurs in about 50% of AD cases and is associated with significantly worse functional outcomes, including increased disability and a faster decline in cognitive abilities.^4^

Shared clinical features and overlapping biological mechanisms,^5^ such as white matter abnormalities,^6^ accelerated brain aging,^7^ synaptic damage,^8^ dopaminergic system dysregulation^9^, and dysregulated acetylcholine system^5^—have increased interest in exploring potential genetic overlaps between schizophrenia and AD. Notably, xanomeline, the active pro-cholinergic component of KarXT (xanomeline-trospium), the newest FDA-approved antipsychotic for schizophrenia, was initially investigated as a potential treatment for Alzheimer’s disease.^10^ Epidemiological evidence indicates a 2.5-fold increased dementia risk in schizophrenia patients,^11^ particularly in late-onset cases (hazard ratio up to 4.22).^12^ However, large-scale GWAS studies initially indicated near-zero common genetic overlap between schizophrenia and Alzheimer’s.^13,14^ Recent polygenic risk score (PRS) analyses offer mixed results, generally finding neutral or inverse correlation between schizophrenia PRS risk and psychotic symptoms in Alzheimer’s.^15-17^ Although recent statistical analyses have begun identifying specific genetic loci associated with both disorders^18^, to date, no studies have examined whether integrating schizophrenia-associated genetic variants could enhance AD risk prediction models or vice versa.

This study explores whether integrating specific schizophrenia-associated single nucleotide polymorphisms (SNPs), identified through statistical criteria from recent PGC GWAS data, enhances the predictive accuracy of Alzheimer’s disease risk models beyond known Alzheimer’s PGC SNPs alone. Recognizing that the genetic risk for complex diseases such as Alzheimer’s likely involves numerous variants with potentially subtle and interactive effects, we leverage machine learning (ML) approaches, which are adept at identifying such complex predictive patterns in high-dimensional data. While PRS approaches integrate a wide spectrum of genetic risk variants, our focused SNP set might highlight more directly implicated genetic markers with stronger, disease-relevant biological effects. We hypothesize that these statistically selected schizophrenia SNPs might capture additional genetic variance relevant to Alzheimer’s pathology or progression, thereby improving predictive models.

## Results

### Overview of Approach

To investigate whether integrating genetic risk associated with schizophrenia (SCZ) could enhance the prediction of Alzheimer’s disease (AD), we utilized summary statistics from the largest available genome-wide association studies (GWAS) for SCZ (PGC3, N≈320k) and AD (PGC-ALZ2, N≈1.1M), alongside whole-genome sequencing data from 7,416 non-Hispanic White individuals from the Alzheimer’s Disease Sequencing Project (ADSP) serving as an independent reference cohort for genotype information. We first curated sets of SNPs reaching the standard genome-wide significance threshold (p ≤ 5×10^−8^) in each GWAS (resulting in N=2,902 AD and N=19,257 SCZ SNPs after initial filtering and harmonization) and subsequently harmonized these variants with the ADSP reference genotype data through standardized procedures, including coordinate conversion (GRCh37 to GRCh38) and allele matching, to ensure data compatibility. To objectively assess the added predictive value of SCZ variants, we employed an iterative feature addition strategy within a machine learning framework, evaluating a selection of distinct classification algorithms capable of capturing different types of predictive patterns. This involved progressively incorporating top-ranked SCZ SNPs—prioritized using a data-driven feature importance metric derived from an initial machine learning model—into a baseline model containing only the AD-associated SNPs. The enhancement in predictive performance was primarily assessed using the standard area under the receiver operating characteristic curve (AUC) metric on a held-out portion of the ADSP data to ensure unbiased evaluation, and the statistical significance of any improvement was rigorously evaluated using multiple established statistical tests. The overall study design and analytical workflow are illustrated in Figure 1.

**Figure 1.**
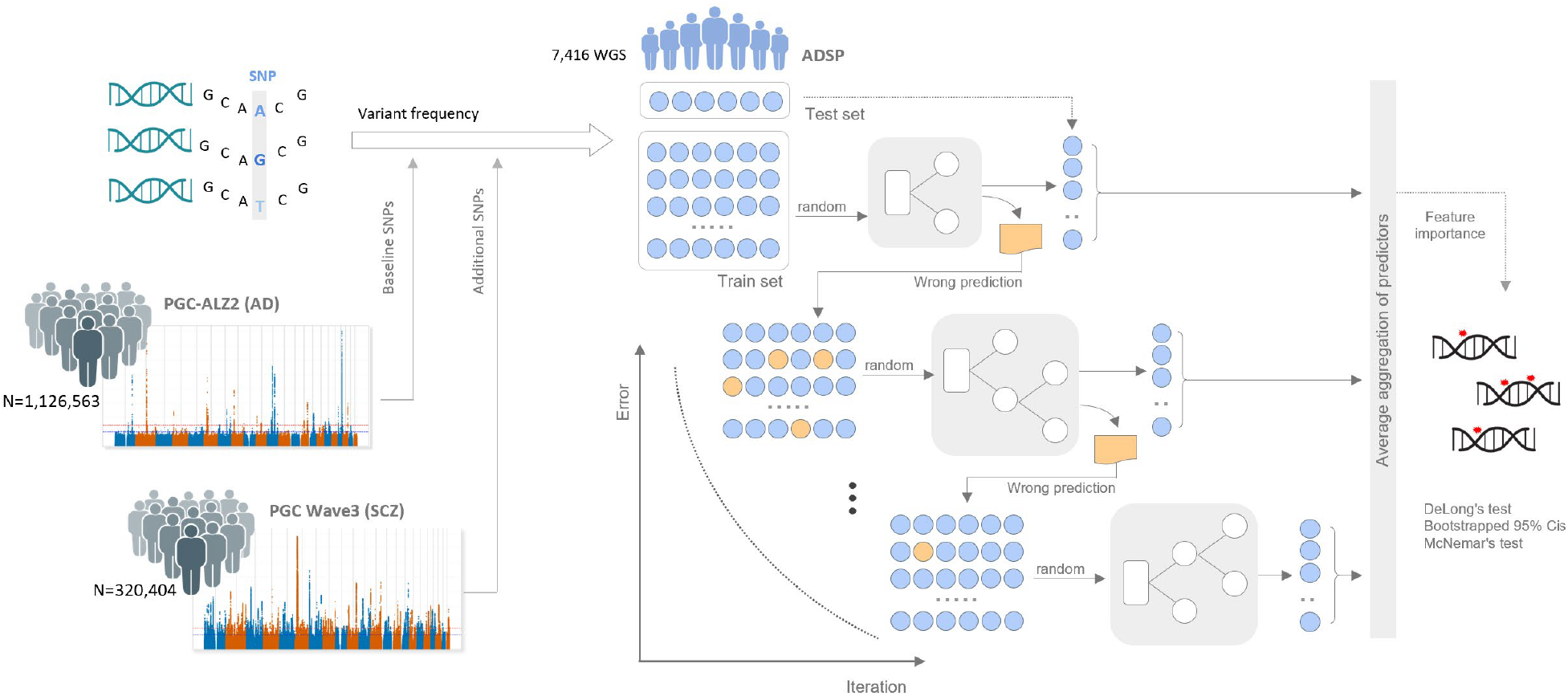
Study workflow for evaluating the predictive value of SCZ SNPs for AD. Genome-wide significant SNPs from large-scale Alzheimer’s disease (AD) and schizophrenia (SCZ) GWAS summary statistics were identified (GRCh37) and subsequently harmonized with reference genotypes from the Alzheimer’s Disease Sequencing Project (ADSP, N=7,416 non-Hispanic Whites, GRCh38) via coordinate conversion and variant matching. An iterative machine learning framework was employed, where top-ranked SCZ SNPs, prioritized using XGBoost feature importance, were progressively added (N=10 to 500) to a baseline model containing only AD-associated SNPs. Performance of three classifiers (Logistic Regression, Random Forest, and XGBoost) was evaluated primarily using the Area Under the Receiver Operating Characteristic Curve (AUC) on held-out test data. Statistical significance of the performance enhancement by the best model (XGBoost with N=50 added SCZ SNPs) compared to its baseline was assessed using DeLong’s test, bootstrapped 95% confidence intervals, and McNemar’s test.

### SNP Data Curation and Harmonization

After filtering for genome-wide significance (p ≤ 5×10^−8^), we identified 3,529 significant AD SNPs and 21,704 significant SCZ SNPs based on GRCh37 coordinates. We converted these coordinates to the GRCh38 assembly using the UCSC liftOver tool. The coordinate conversion process successfully mapped the vast majority of variants; mapping failures were limited to 11 AD SNPs, comprising 0.31 percent of the initial AD set, and 8 SCZ SNPs, comprising 0.04 percent of the initial SCZ set. Following the exclusion of these unmapped SNPs, subsequent matching against the ADSP reference data yielded our final SNP sets, which consisted of 2,902 AD-associated SNPs and 19,257 SCZ-associated SNPs for analysis in our sample of 7,416 non-Hispanic White individuals from the ADSP dataset. Of these curated SNPs, 21 were shared between the AD and SCZ genome-wide significant lists.

### SCZ SNP integration enhances AD prediction

Integrating SCZ-associated variants with AD SNPs generally improved predictive performance across the three machine learning models tested: XGBoost, RF, and LR. Baseline AUC values, using only AD SNPs (N=0), ranged from 0.5732 (LR) to 0.6032 (XGBoost) (Table 1). Optimal AUC performance for each model was typically achieved with a relatively small number of added SCZ SNPs (N=10 for LR, N=40 for RF, N=50 for XGBoost). The XGBoost classifier achieved the highest overall performance, reaching a peak AUC of 0.6400 when incorporating the top 50 SCZ SNPs. This represents a 0.0368 improvement over its baseline performance (Table 1). This AUC improvement for XGBoost (N=50 vs. N=0) was statistically significant based on DeLong’s test (p = 0.0005). Further supporting the enhanced performance of the best model (XGBoost, N=50), bootstrapped 95% confidence intervals (CIs) for all evaluated metrics consistently improved compared to the baseline model (Table 2). Notably, the AUC 95% CI shifted from (0.5745– 0.6335) for the baseline to (0.6124–0.6688) for the best model, exhibiting minimal overlap. Similar improvements in CIs were observed for Accuracy, F1 Score, Precision, Recall, and Specificity (Table 2). Additionally, McNemar’s test indicated a significant difference in classification predictions between the baseline and the best XGBoost model (p = 0.0018).

**Table 1.** Comparison of Model Performance Integrating SCZ SNPs.

| Model | Baseline AUC (N=0) | Best N | Best AUC | AUC at Max N (500) | Improvement over Baseline |
| --- | --- | --- | --- | --- | --- |
| XGBoost | 0.6032 | 50 | 0.64 | 0.6055 | 0.0368 |
| Random Forest | 0.5958 | 40 | 0.6063 | 0.5942 | 0.0105 |
| Logistic Regression | 0.5732 | 10 | 0.5751 | 0.5748 | 0.0018 |

**Table 2.** Performance Metrics Comparison: Baseline vs. Best Model (XGBoost, N=50 SCZ SNPs)

| Metric | Baseline Value | Baseline CI Lower | Baseline CI Upper | BestN Value | BestN CI Lower | BestN CI Upper |
| --- | --- | --- | --- | --- | --- | --- |
| AUC | 0.6032 | 0.5745 | 0.6335 | 0.64 | 0.6124 | 0.6688 |
| Accuracy | 0.5822 | 0.5566 | 0.6085 | 0.6199 | 0.5963 | 0.6429 |
| F1 Score | 0.663 | 0.6373 | 0.6876 | 0.6921 | 0.6674 | 0.7153 |
| Precision | 0.6187 | 0.5892 | 0.6492 | 0.6483 | 0.6175 | 0.6772 |
| Sensitivity | 0.7143 | 0.6828 | 0.7444 | 0.7424 | 0.7123 | 0.7725 |
| Specificity | 0.4032 | 0.3654 | 0.4418 | 0.454 | 0.4168 | 0.493 |

As detailed in Table 1, Random Forest performance peaked at N=40 (AUC=0.6063), while Logistic Regression showed only modest improvement, peaking at N=10 (AUC=0.5751). Performance for most models did not substantially increase when adding a larger number of SCZ SNPs (e.g., N=500). This suggests that only specific SCZ-associated variants contribute meaningfully to AD prediction. Detailed performance metrics across all tested N values for AUC, Accuracy, and F1 Score are available in the Supplementary Material (Table S1).

### Identification of influential SCZ SNPs

We identified the top SCZ SNPs that contributed most significantly to AD prediction performance using the XGBoost feature importance metric. Table 3 lists the top 50 contributing variants, including the three most influential: rs139779037, rs9257331, and rs7815859.

**Table 3.**
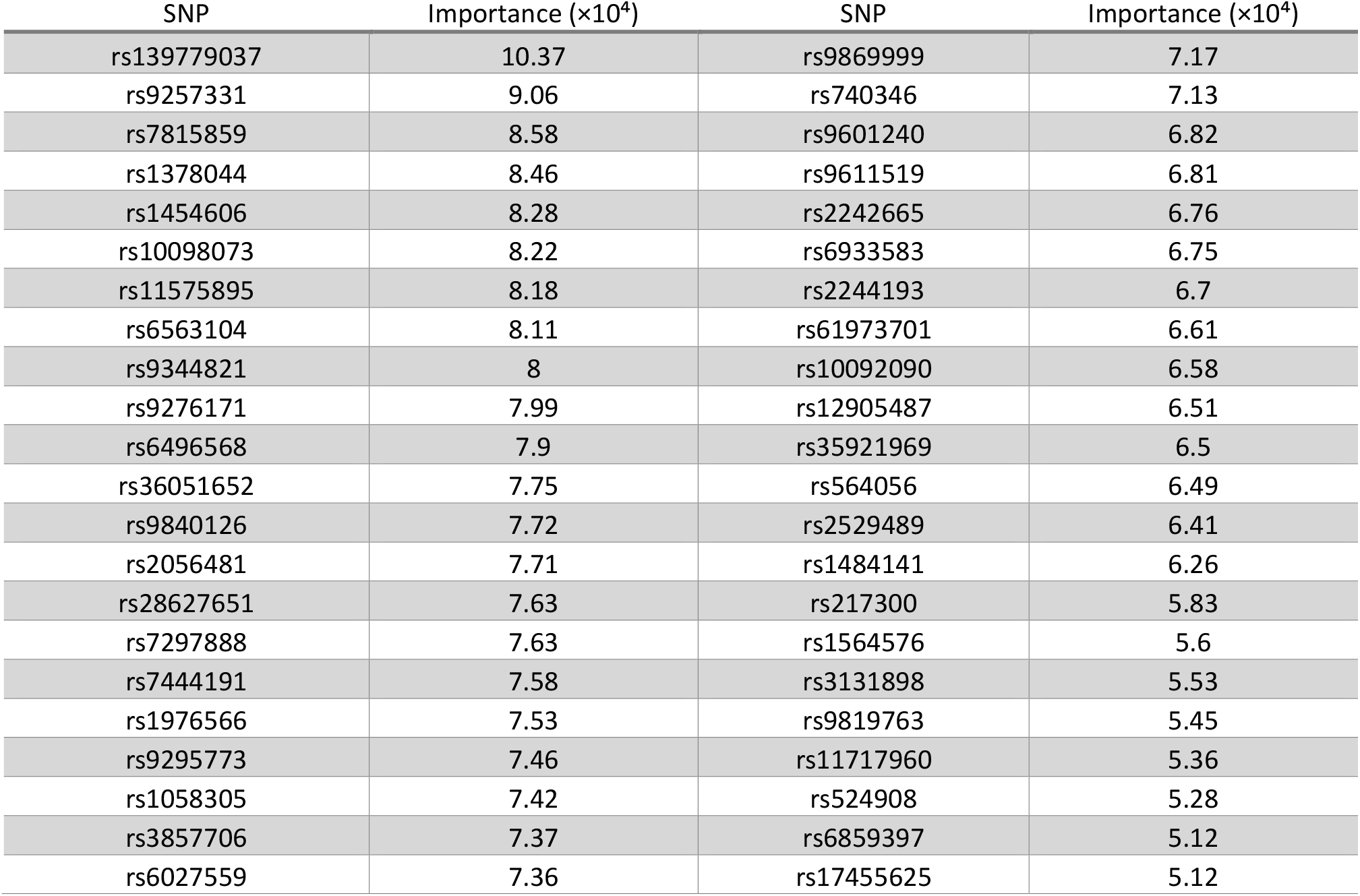
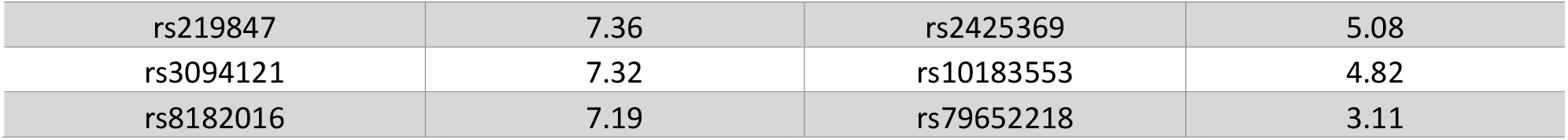
SCZ SNPs ranking.

### Comparative ROC curve analysis

Comparative ROC curve analysis (Figure S1) confirmed that the optimal combined feature set (AD SNPs + top 50 SCZ SNPs) consistently outperformed both the AD-only and SCZ-only models across the entire range of sensitivity and specificity thresholds when evaluated using XGBoost, which demonstrated the highest overall performance.

## Discussion

This study investigated whether incorporating schizophrenia-associated genetic variants could enhance AD risk prediction models. The inclusion of the top 50 schizophrenia-associated SNPs, yielded a significant, albeit modest, improvement in AD predictive accuracy (AUC increased from 0.603 to 0.640, p=0.0005). This finding supports meaningful genetic overlap and pleiotropy between the disorders, despite minimal direct overlap (N=21 SNPs) observed between the initial genome-wide significant SNP sets for AD and SCZ. Furthermore, the predictive enhancement appears to stem largely from novel genetic factors from an AD perspective, as none of the top 50 prioritized SCZ SNPs were found within the curated significant AD SNP set. This reveals previously unrecognized genetic contributions to AD risk beyond established loci like APOE, BIN1, and TREM2.^19^

Several identified schizophrenia-associated SNPs implicate biological pathways already recognized in AD, including immune regulation, neuroinflammation, tau protein and microtubule dynamics, and synaptic vesicle trafficking. Genetic variants within the Major Histocompatibility Complex (MHC) region, such as rs9257331, rs9276171, rs3094121, rs3131898, support shared immunological mechanisms involving abnormal complement-mediated synaptic pruning, linking complement component 4 in schizophrenia^20^ and complement components C1q/C3 in AD.^21,22^ In addition, the SNP rs11575895 within the tau-encoding MAPT gene (chromosome 17q21.31), highlights shared tau-related pathologies between schizophrenia and AD. Tau pathology, classically associated with AD,^23,24^ is increasingly recognized in schizophrenia concerning microtubule stability and axonal transport.^25,26^ This SNP is further recognized as a common risk variant for both schizophrenia and executive function traits, indicative of its pleiotropic neurodevelopmental role.^27^

Beyond established pathways, this study also identified novel schizophrenia-associated SNPs with previously unexplored connections to AD, including variants in genes central to synaptic transmission such as TSNARE1^28^ (rs10098073, rs10092090) and SNAP91 (rs217300).^29^ These genes regulate essential synaptic vesicle trafficking processes (exocytosis and clathrin-mediated endocytosis, respectively) that are disrupted early in AD.^30^ While TSNARE1 has previously been indirectly linked to AD through epigenetic mechanisms,^31^ these findings highlight direct genetic associations warranting further exploration. Similarly, although related genes such as BIN1 and PICALM are established in AD loci,^32^ SNAP91 represents a relatively unexplored genetic component, further implicating synaptic vesicle recycling as a potentially critical yet underexplored pathway in AD.

In addition, the analysis uncovered novel schizophrenia-associated SNPs that were not previously implicated in AD, and whose roles in disease pathology remain unclear. These SNPs do not align with established schizophrenia risk genes (e.g., CACNA1C,^33-35^ ZNF804A,^36^ MIR137,^37^ DRD2,^38^ TCF4,^39^ NRGN^39^)^14^ and also lack clear connections to known AD pathways. Their identification raises important questions. These novel variants might reflect previously unrecognized biological process contributing to both disorders, or alternatively; they might represent subtle genetic effects arising from methodological variability or the highly polygenic nature of schizophrenia. Further research will be necessary to clarify their relevance to AD risk and to understand their biological implications.

There are several limitations. First, our selection of schizophrenia-associated SNPs was based exclusively on statistical significance criteria from existing GWAS data, potentially leading to false positives or inflated effect sizes and excluding biologically relevant variants below the significance threshold. Thus, the actual genetic overlap between AD and schizophrenia might be broader than captured here. Second, although statistically significant, the observed improvement in predictive accuracy remains modest, reflecting that additional genetic, epigenetic, environmental, and lifestyle factors influencing AD risk were not included. Third, we analyzed only additive effects of common variants, omitting potential gene–gene interactions and rare variants that could further inform shared disease mechanisms. Furthermore, our analysis was restricted to individuals of non-Hispanic White ancestry from the ADSP dataset. Genetic architectures and risk variants for both SCZ and AD can differ across populations, thus limiting the generalizability of our current findings. Finally, replication in independent, diverse cohorts and functional genomic studies are required to confirm the robustness and biological significance of these genetic associations.

Despite these limitations, our findings demonstrate the utility of cross-disorder genetic analyses in complex neuropsychiatric diseases. Incorporating schizophrenia-associated genetic variants into AD prediction models produced a statistically significant, albeit modest, improvement in prediction accuracy. This approach successfully identified shared biological mechanisms involving immune-mediated synaptic pruning, tau-related cytoskeletal dysfunction, and synaptic vesicle trafficking. These findings strongly support existing evidence for shared molecular pathways between schizophrenia and AD and provide novel genetic targets for future cross-disorder research. Future studies should aim to validate these associations in larger cohorts, investigate their functional implications in neuronal and synaptic pathology, and explore reciprocal applications of integrating Alzheimer’s variants into schizophrenia risk models. Ultimately, leveraging cross-disorder genetic analyses promises to deepen our understanding of complex neuropsychiatric disorders, enhance predictive models, and guide therapeutic development.

## Method

### Data Sources and Preprocessing

We utilized summary statistics from two large-scale genome-wide association studies (GWAS): the Alzheimer’s Disease (AD) study by Wightman et al. (2021; N=1,126,563; PGC-ALZ2) ^40^ and the Schizophrenia (SCZ) study by Trubetskoy et al. (2022; N = 320,404; PGC wave 3) ^41^. Both datasets were based on the GRCh37 reference genome. These studies employed rigorous quality control (QC), including standard sample-level and SNP-level filtering based on call rates, heterozygosity, sex concordance, population structure, Hardy-Weinberg equilibrium (typically P > 1×10^−6^ in controls), imputation quality (e.g., INFO > 0.3), and minor allele frequency (typically > 0.1–1%). From these datasets, we selected SNPs reaching genome-wide significance (P ≤ 5×10^−8^), ensuring valid coordinates and P-values, and removed positional duplicates.

For genotype reference data, we used whole genome sequencing (WGS) data from 7,416 non-Hispanic White individuals from the Alzheimer’s Disease Sequencing Project (ADSP). This dataset underwent QC using standard filters (SNP missingness < 0.05, MAF ≥ 0.01, individual missingness < 0.05). As the ADSP data utilized the GRCh38 assembly, we converted the coordinates of the selected GWAS SNPs from GRCh37 to GRCh38 using the UCSC liftOver tool^42^ with the hg19-to-hg38 chain file. SNPs successfully mapped to GRCh38 were subsequently filtered by matching against variants present in the ADSP reference data, yielding the final AD and SCZ SNP sets used for analysis.

### Machine Learning Framework

We employed an iterative feature addition framework to assess the potential of SCZ-associated SNPs to enhance AD prediction models. Data preparation involved extracting genotypes for the selected AD and SCZ SNP sets from the ADSP WGS data using PLINK v1.9^20^. The AD case/control status served as the binary phenotype (0=control, 1=AD). Missing genotype values were imputed using mean imputation implemented via scikit-learn^43^.

To prioritize SCZ variants, an XGBoost classifier^44^ was trained using only the SCZ-associated SNPs, and SNPs were ranked based on the resulting feature importance scores. A baseline model was established using only AD SNPs. Subsequently, models were trained by progressively adding the top N ranked SCZ SNPs (N = 10, 20, 30, …, 500) to the baseline AD SNP set. For each N, the ADSP dataset was stratified by phenotype and split into training (80%) and testing (20%) sets for model evaluation.

We evaluated three classification algorithms: Logistic Regression (with L2 regularization, balanced class weights, and preceding feature standardization), Random Forest (with balanced class weights), and XGBoost. Model performance was assessed on the test set using the area under the receiver operating characteristic curve (AUC), accuracy, F1-score, precision, recall, and specificity.

Feature importance scores for the added SCZ SNPs were extracted from the trained XGBoost models at each N step to evaluate their contribution within the combined feature set. All analyses were performed using Python 3. Key libraries included pandas^45^, NumPy^46^, scikit-learn, XGBoost, Matplotlib^47^, MLstatkit, and statsmodels.

### Statistical Significance Testing

To rigorously evaluate performance improvements, the best-performing model configuration (algorithm and N value yielding the highest test set AUC) was compared against its corresponding baseline model (N=0) using three statistical tests on the held-out test set predictions. First, DeLong’s test for correlated ROC curves was used to compare AUC values. Second, 95% confidence intervals (CIs) for AUC, accuracy, F1-score, precision, recall, and specificity were calculated using bootstrapping (1000 resamples) to assess the range and overlap of performance estimates. Third, McNemar’s test was employed to evaluate systematic differences in classification predictions between the baseline and best models.

## Data availability

The summary statistics utilized in this study are publicly available. Alzheimer’s disease GWAS summary statistics from Wightman et al. (2021; PGC-ALZ2) and Schizophrenia GWAS summary statistics from Trubetskoy et al. (2022; PGC wave 3) can be accessed through the Psychiatric Genomics Consortium (PGC) data download portal (https://pgc.unc.edu/for-researchers/download-results/).

The Alzheimer’s Disease Sequencing Project (ADSP) whole-genome sequencing (WGS) data used as reference genotypes are available through the NIH National Institute on Aging Genetics of Alzheimer’s Disease Data Storage Site (NIAGADS) under accession number NG00067 (https://dss.niagads.org/datasets/ng00067/). Access to individual-level ADSP WGS data requires application and approval by the NIAGADS Data Access Committee (DAC). Qualified investigators can submit a Data Access Request via the NIAGADS Data Sharing Service (DSS) portal. More information about the ADSP datasets and application procedures can be found on the NIAGADS website (https://dss.niagads.org/).

## Code availability

The analysis code and result files supporting the findings of this study are available on GitHub: https://github.com/taehojo/AD-SCZ

## Acknowledgements

We thank the participants of the studies contributing to the PGC-ALZ2 and PGC3 GWAS datasets, and the participants of the Alzheimer’s Disease Sequencing Project (ADSP). We acknowledge the Psychiatric Genomics Consortium (PGC) for access to the GWAS summary statistics. We acknowledge the ADSP consortia for generating and providing the WGS data used as a reference panel in this study.

## Author information

These authors contributed equally: Taeho Jo, Carol S. Lim

### Contributions

T.J. and C.S.L. contributed equally to the conceptualization and design of the study. T.J. developed the methodology, performed data curation and machine-learning analysis, and prepared visualization. C.S.L. provided interpretation of results and formulated clinical insights, including genotype-phenotype associations. Both authors contributed to drafting, revising, and finalizing the manuscript, and approved the final version.

## Supplementary information

Supplementary Information

